# Greater recalled pain and movement-evoked pain are associated with longer 400-meter walk and repeat stair climb time: the Study of Muscle, Mobility and Aging

**DOI:** 10.1101/2024.10.19.24315822

**Authors:** Theresa Mau, Haley N. Barnes, Corey B. Simon, Megan Hetherington-Rauth, Scott R. Bauer, Elsa S. Strotmeyer, Nancy E. Lane, Michael C. Rowbotham, Ashley A. Weaver, Terri L. Blackwell, Yurun Cai, Nancy W. Glynn, Bret H. Goodpaster, Steven R. Cummings, Anne B. Newman, Peggy M. Cawthon, Stephen B. Kritchevsky

## Abstract

**Background:** Musculoskeletal pain frequently accompanies the development of mobility disability and falls in old age. To better understand this, we aimed to quantify the impact of different pain measures—recalled pain and movement-evoked pain—on 400-meter walk and stair climb time in older adults participating in the Study of Muscle, Mobility and Aging (SOMMA).

**Methods:** In SOMMA (N=879, age=76.3 ± 5.0 years, 59% women, 84% Non-Hispanic White), participants completed usual pace 400m walk (avg=6.6 ± 1.2 min.) and repeat stair climb tests (avg=26.6 ± 7.2 sec.). Assessments of recalled pain included the Brief Pain Inventory short form (BPI-*sf*), total lower body pain (lower back, hips, knees, feet/ankles), stiffness (hip or knee), and Neuropathy Total Symptom Score (NTSS-6). Movement-evoked pain was assessed separately before and after the 400m walk and repeat stair climb tasks. Multivariable linear regression modeled the associations of pain with time to complete the tasks, reported as β[95%CI] expressed per SD increment of pain measure or β[95%CI] per pain categories, adjusted for age, sex, race, ethnicity, body mass index, prescription medications, and depressive symptoms.

**Results:** Greater degree of any pain measure was associated with longer physical performance time, though intercorrelations between recalled pain measures varied (r=0.13-0.57, *p*<0.05 for all). For each SD increment in lower body pain, participants had longer walk time (by 10.5 sec [6.1, 14.8]) and stair climb (by 0.6 sec [0.1, 1.1]). Compared to participants with no change in pain upon movement, walk time was longer in those with more pain upon movement (19.5 sec [10.3, 28.7]) (*p*<0.001) but not those with less pain upon movement; stair climb showed similar patterns.

**Conclusions:** Recalled and movement-evoked pain measures were weakly correlated with one another but similarly associated with time to complete 400m walk and stair climb tests. Different pain assessments capture different functional domains of pain but have similar associations with physical performance in these older adults.

## Introduction

Pain is a very common symptom in older adults. Nationally representative data suggest that nearly 75% of older adults experience chronic multisite pain.^1^ Pain is especially prevalent among older women and in those with musculoskeletal conditions, obesity, or mood disorders.^1^ The negative impact of pain on healthy aging is evident: many different measures of pain have been associated with lower and upper extremity disability.^2^ Pain may also directly cause, or be perceived to cause, mobility disability. In at least one study, lower back pain is the primary symptom older adults attribute to their difficulty in walking.^3^ Pain also predicts poor outcomes, such as falls, in older adults.^4–7^ While the relationship between chronic musculoskeletal pain and disability in older age is established, pain remains complex in its influences and manifestations.

Many measures of pain include features of both nociceptive and neuropathic pain.^8^ Musculoskeletal pain depends on nociceptors, specialized nerve endings in muscle, to translate tissue and other damage to the central nervous system as nociceptive pain.^9^ Pain signaling relies on the nervous system; damage or inflammation of the nerve and other parts of the nervous system are key stimuli in neuropathic pain, another category of pain. More recently, a third broad category of pain—nociplastic—was described^8^ to recognize complex pain syndromes that are widespread (i.e., afflicting pain across multiple regions of the body) and occur in the absence of tissue aberration or damage. Fibromyalgia is an example of nociplastic pain. However, pain influences are multi-factorial, and it remains difficult to disentangle what types of pain are captured in pain assessments collected in clinical and research settings. Furthermore, it is unclear if relationships to functional outcomes are consistent across the multiplicity of pain measures, or if specific pain measures are especially strong predictors of poor functioning.

When pain is assessed clinically, patients are typically asked to rate resting pain while sitting in an exam room. On the other hand, in research settings, pain is often assessed via questionnaires that rely on people to recall pain in the previous hours to years and may include pain experienced while resting, during movement or physical activity. These recall assessments range from tallying pain anatomic sites to rating pain severity and frequency. One such commonly used assessment is the Brief Pain Inventory short form (BPI-*sf*)^10^ which counts the pain locations and also queries global pain severity (past, current, worst, average), but the severity rating is not specific to any pain site. Another pain assessment tool is the Neuropathy Total Symptom Score 6-item (NTSS-6) which queries neuropathic pain sensory symptom (e.g., numbness, insensitivity, burning pain) severity and frequency below the knees.^11^ In contrast, pain assessments within osteoarthritis studies focus on localized sites of musculoskeletal pain and stiffness, and these questions include recall pain and stiffness experienced with movement or physical activities.^12^ Measuring pain concurrently with validated physical performance tests— movement-evoked pain—could help identify pain that specifically disrupts function.^13^ Few studies have incorporated and quantified the impact of different measures of pain, including both recalled and movement-evoked, on physical performance outcomes in older adults.

We have recently shown in the Study of Muscle, Mobility and Aging (SOMMA) that musculoskeletal pain and stiffness, amongst dozens of clinical and physiologic characteristics, were major factors associated with the walk speed of older adults.^14^ This current report aimed to understand how measures of recalled pain (resting and during activity) and movement-evoked pain (during the 400m walk and stair climb) relate to objective measures of physical performance, the 400m walk and repeat stair climb test, in older adults.

## Methods

### Study cohort

The Study of Muscle, Mobility and Aging (SOMMA) is a longitudinal cohort study of the biology of human aging, with a particular focus on mobility. Details on the study design have been published.^15^ Baseline assessments between April 2019 and December 2021 were completed in 879 adults aged 70 or older at the University of Pittsburgh and Wake Forest University School of Medicine. Individuals ≥70 years old were eligible to participate if willing and able to complete a skeletal muscle biopsy and undergo magnetic resonance (MR) spectroscopy. Individuals were excluded if they had body mass index (BMI) ≥40 kg/m^2^, had an active malignancy or dementia, or any medical contraindication to muscle biopsy or MR. Eligibility also included participant ability to complete the 400-meter (400m) walk within 15 minutes. Individuals who appeared as they might not be able to complete the 400m walk had to complete a short distance walk (4 meters) at the in-person screening baseline visit to ensure that their walk speed was ≥0.6 m/s. If a participant felt they could not safely attempt the 400m walk without an assistive device other than a straight cane, they were considered ineligible to enroll for SOMMA. SOMMA was approved by the Western IRB-Copernicus Group Institutional Review Board (WCG-IRB; 20180764). All participants provided written informed consent.

### Clinic characteristics and prescription medications

Participants completed questionnaires and exams over three clinic visits—which were conducted within six to eight weeks of each other. Participants reported date of birth, sex, race and ethnicity, education, alcohol intake (drinks/week), smoking history (no, past, current), and medical history. The 10-item version of the Center of Epidemiologic Studies Depression Scale (CESD-10) was used to determine depressive symptoms (score≥10, range: 0-30).^16^ A subset of participants (N=658) had radiographic scores on knee osteoarthritis (OA) which were collected ∼1 year after the baseline exam on knee radiographs.^17^ The presence of knee OA was defined as Kellgren-Lawrence (KLG) scores ≥2. Bilateral knee OA was defined as having KLG ≥2 in both left and right sides in any combination of the tibio-femoral and patella-femoral sites.^18^ Participants were asked to bring all prescription medications they had taken within 30 days of the first clinic visit. All medications were entered into an electronic database, verified by pill bottle examination. Each medication was matched to a medication code and generic ingredient name(s) based on RxTerms^19^, a standardized nomenclature for clinical drugs (National Library of Medicine, Bethesda, MD). Using RxMix^20^, these medication codes were then matched to the WHODrug Anatomical Therapeutic Chemical (ATC) classification system (Uppsala Monitoring Centre, Uppsala, Sweden).^21^ The total count of prescription medications were summed and specific prescription analgesic medications were coded into the following groups: antidepressants (non-selective monoamine reuptake/oxidase inhibitors, selective serotonin reuptake inhibitors), anti-inflammatories (acetic acid derivatives and related substances, oxicams, propionic acid derivatives, coxibs), and opioids (natural opium alkaloids, opioids in combination with non-opioid analgesics).

### Overview of pain assessments

Participants completed pain assessments (**Supplementary Table 1**) at baseline that can be broadly grouped into two sets of measures—recalled pain and movement-evoked pain: **1**) Questionnaires on different measures of *recalled pain* include the Brief Pain Inventory short form (BPI-*sf*), lower body pain (lower back, hips, knees, feet/ankles assessed separately), hip/knee stiffness, and Neuropathy Total Symptom Score 6-item version (NTSS-6). **2**) *Movement-evoked pain* is assessed concurrently to the 400m walk and stair climb tests, two separate pain scores are calculated for each physical performance test as described below.

### Recalled pain from the Brief Pain Inventory-short form (BPI-sf): Global pain severity

A baseline clinic questionnaire included the BPI-*sf* which assesses recalled pain (0 to 10 scale) with questions on the worst pain in the last 24 hours, least pain in the last 24 hours, pain on average, and current pain.^10^ These four ratings were averaged to generate the global pain severity score.^22^ Cut points were used to categorize participants by global pain severity groups: none (x=0), mild (0<x<2), moderate (2≤x<4), moderate-to-severe (4≤x≤10) pain.^2^ Participants were queried for any presence of pain at 22 different anatomic sites (includes laterality—e.g., left versus right shoulder pain counts as 2 sites). The number of pain sites were used to classify participant pain site burden into four mutually exclusive categories: none, single site, multisite (2+ but not widespread), and widespread pain. Based on the American College of Rheumatology (ACR) guidelines^23^, widespread pain can be identified if these three conditions are met: there is pain above and below the waist, pain on the left and right side of the body, and axial skeletal pain (back or non-anginal chest pain). Self-reported medical history of atrial fibrillation, aortic stenosis, heart failure, and heart attack or myocardial infarction allowed us to exclude participants with potential anginal chest pain (and no back pain) from the widespread pain category.

### Recalled pain in specific anatomic locations: lower body pain and hip/knee stiffness

Self-assessment questionnaires included questions on chronic pain in four lower body anatomic locations: knees, hips, feet or ankles, and lower back. Participants were queried about pain frequency (never, monthly, weekly, daily, always) and pain severity (none, mild, moderate, severe, extreme) in each anatomic site while walking on flat surface, going up or down stairs, and laterality of pain. All pain frequency and severity questions were summed to generate a separate score for each anatomic location (knees, hip, feet/ankles, or lower back). A total lower body pain score (51 points possible) was calculated by summing the four sub scores at each anatomic location. Stiffness in the hip or knee (left and right) were also assessed by questionnaire. The severity of stiffness after first waking up, after resting later in the day, current stiffness, frequency of how often it was difficult to walk due to stiffness in past week, and number of locations were scored to generate a hip/knee stiffness score (19 points possible).

### Recalled pain by Neuropathy Total Symptom Score 6-item (NTSS-6): somatosensory pain

Peripheral nerve pain symptoms (numbness, prickling, allodynia, lancinating, burning, and aching) were assessed with the Neuropathy Total Symptom Score, 6-item version (NTSS-6).^11^ If participants report experiencing the sensation within the past 24 hours, then questions further queried the sensation frequency (occasional, often, continuous) and intensity (mild, moderate, severe). Total somatosensory pain scores across the six nerve pain sensations were calculated using a validated, weighted scale that accounts for the interaction of symptom intensity and frequency (21.96 points possible).^11^ SOMMA received permission to use this scale (CC BY-ND v4.0 International License).

### 400m walk test and movement-evoked pain assessment for 400m walk test

The 400m walk was assessed at the participant’s usual (i.e., preferred) pace for 10 laps on a 40-meter course (20m space marked by cones in a long corridor) with a straight cane if necessary (N=24).^24,25^ Time to complete the 400m walk (sec) included any standing rests (up to 1 min.) taken by the participant during the test. All participants completed the test as a requirement for including in SOMMA. Before, during (immediately after lap 4), and after the 400m walk test, participants rated their current pain on a scale of 0 to 10 where 0 is no pain and 10 is the worst pain imaginable. Movement-evoked pain (aggregate score) was calculated by subtracting baseline pain rating prior to the test from the summed pain score at lap 4 and after test (possible score range –10 to 10).^13^ Those with negative, neutral/zero, and positive aggregate scores are categorized as less pain upon movement, no change, and more pain upon movement.

### Repeat stair climb test and movement-evoked pain assessment for stair climb test

A set of four standard stairs of steps (10-inch depth and 6-inch height) were used for the timed, repeated 3-lap, 4-step stair climb test (sec). Participants ascended and descended the four stairs for three consecutive cycles (N=845) with a handrail for assistance if necessary, over a third of the participants used the handrail (N=353, 42%).^13^ The cumulative time of the three laps included only completed laps which consisted of the ascend and descend split times from base to the top step that was recorded for each lap. A total of 34 participants were missing repeat stair climb data: 21 due to clinic or equipment limitations and 13 due to safety concerns or incomplete testing data. Before the repeat stair climb test, participants rated current pain on a scale of 0 to 10 where 0 is no pain and 10 is the worst pain imaginable. After the test, participants rated their worst pain experienced during or after the stair climb test. Participant movement-evoked pain (delta score) was calculated by subtracting the baseline pain rating (prior to the test) from the peak pain reported after stair climb test (possible score range –10 to 10). Those with negative, neutral/zero, and positive delta scores are categorized as less pain upon movement, no change, and more pain upon movement.

## Statistical Methods

For baseline characteristics, *p* for linear trend across BPI-*sf* global pain severity categories (none, mild, moderate, moderate-to-severe) was calculated using linear regression for normally distributed continuous variables and Jonckheere-Terpstra test for skewed or categorical variables. Spearman correlation was used to assess the relationship between continuous pain measures and other covariates.

We examined the associations between numerous pain measures with two physical performance outcomes: time to complete 400m walk and repeat stair climb time. Results for categorical predictors such as pain site classification (i.e., no pain site, single pain site, multisite pain, widespread pain), somatosensory pain (NTSS-6 score; none, mild, moderate-to-severe), movement-evoked pain for 400m walk (aggregate score; less pain upon movement, no change, more pain), and movement-evoked pain for stair climb (delta score; less pain upon movement, no change, more pain) are expressed as beta coefficients (95% CI) in relation to the referent group/category. Global pain severity, total lower body pain (lower back, hips, knees, feet/ankles), and hip/knee stiffness are treated as continuous variables, with results expressed as beta coefficients (β) with 95% confidence intervals (CI) per 1 SD increment in pain measure. We also performed models for anatomic site-specific lower body pain, first for each site individually and then for all anatomic sites in the same model. Model 1 (minimally adjusted) adjustments included site, age, sex, race/ethnicity (non-Hispanic White vs. Other), and BMI. Model 2 further adjusted for number of prescription medications taken in past 30 days and depressive symptoms (CESD-10). We examined whether associations between pain and physical performance (400m walk and stair climb time) varied by sex in linear regression models including site, the pain variable of interest, sex, and their product (pain variable*sex). Interaction models were also run to evaluate whether associations between pain and physical performance varied by presence of knee OA. Additional interaction models evaluated whether the association varied by knee OA classification (i.e., no knee OA, unilateral knee OA, bilateral knee OA). To test the association between global pain severity and physical performance, linear regression models were stratified by classification. Analyses were completed in SAS version 9.4 and R version 4.3.

## Results

### Analytic cohort and baseline characteristics

All participants (N=879) completed at least one of the following recalled pain assessments: global pain severity (BPI-*sf*, N=879), lower body pain and stiffness (N=876), and somatosensory pain symptoms (NTSS-6, N=866). No participants had missing data for the 400m walk test, and 34 had missing data for the repeat stair climb test. All these participants have completed the corresponding movement-evoked pain assessments (400m walk, N=879; stair climb, N=845). Terminology of pain measures by assessment is summarized in **Supplementary Table 1**.

At SOMMA baseline, 32% (N=279) reported no pain, 36% (N=318) a single pain site, 24% (N=213) multisite pain, and 8% (N=69) met the definitions for widespread pain. The most common anatomic pain sites were lower back, knees, shoulders, and hips (**Supplementary Table 2**). People with the greatest pain severity were more likely to be women, and they had a higher BMI, a lower level of education, greater depressive symptomology, and took a greater number of prescription medications compared to groups with lower pain severity (**Table 1**). Besides self-reported arthritis, there were no significant trends (not shown) in pain severity with other individual conditions in the multimorbidity index. All other measures of recalled pain and movement-evoked pain were summarized across the categories of global pain severity.

**Table 1.**
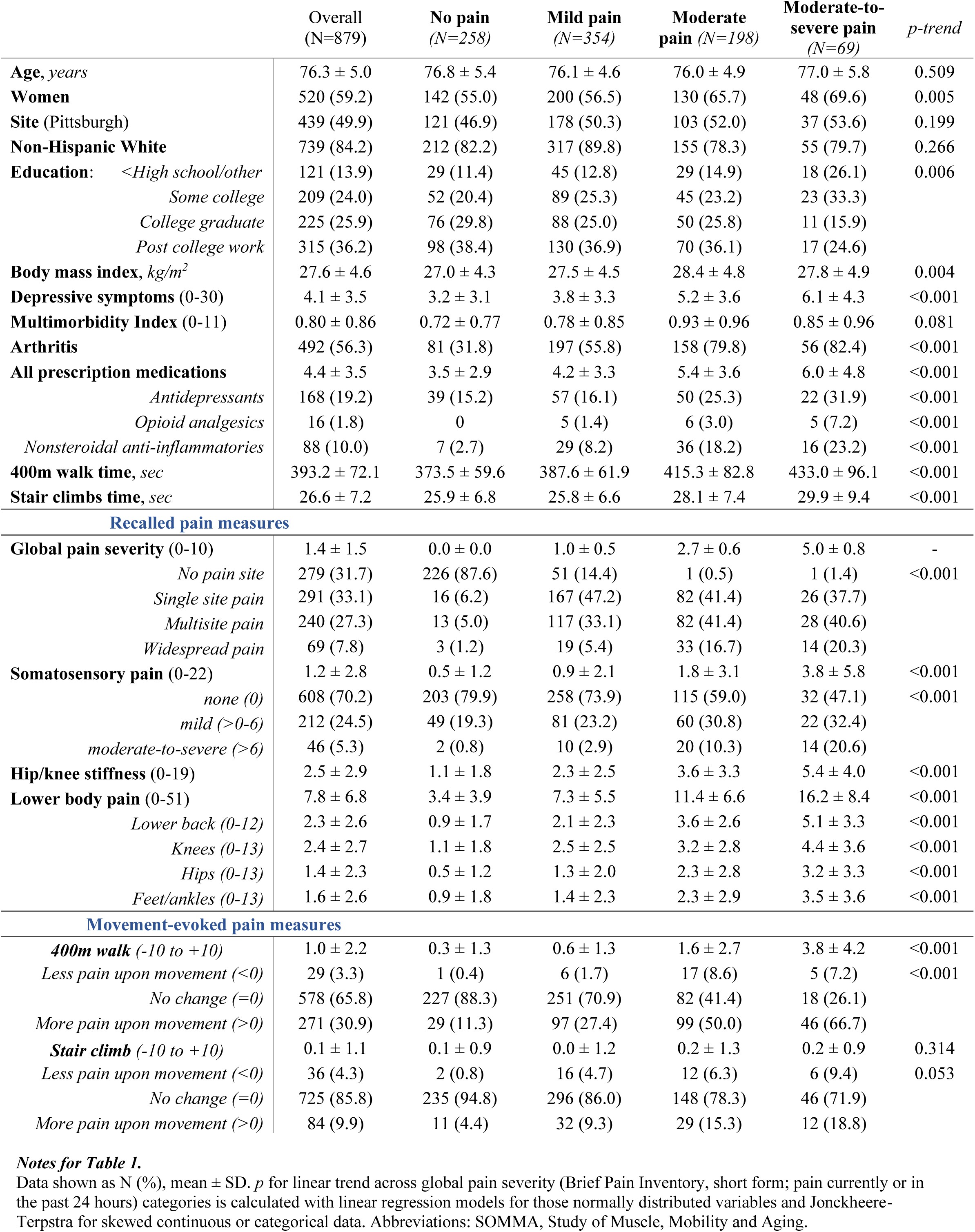
Baseline characteristics of SOMMA older adults (≥70 years) by global pain severity.

|  | Overall<br>(N=879) | No pain<br>(N=258) | Mild pain<br>(N=354) | Moderate<br>pain (N=198) | Moderate-to-severe pain<br>(N=69) | *p-trend* |
| --- | --- | --- | --- | --- | --- | --- |
| **Age, years** | 76.3 ± 5.0 | 76.8 ± 5.4 | 76.1 ± 4.6 | 76.0 ± 4.9 | 77.0 ± 5.8 | 0.509 |
| **Women** | 520 (59.2) | 142 (55.0) | 200 (56.5) | 130 (65.7) | 48 (69.6) | 0.005 |
| **Site (Pittsburgh)** | 439 (49.9) | 121 (46.9) | 178 (50.3) | 103 (52.0) | 37 (53.6) | 0.199 |
| **Non-Hispanic White** | 739 (84.2) | 212 (82.2) | 317 (89.8) | 155 (78.3) | 55 (79.7) | 0.266 |
| **Education:** <High school/other | 121 (13.9) | 29 (11.4) | 45 (12.8) | 29 (14.9) | 18 (26.1) | 0.006 |
| Some college | 209 (24.0) | 52 (20.4) | 89 (25.3) | 45 (23.2) | 23 (33.3) |  |
| College graduate | 225 (25.9) | 76 (29.8) | 88 (25.0) | 50 (25.8) | 11 (15.9) |  |
| Post college work | 315 (36.2) | 98 (38.4) | 130 (36.9) | 70 (36.1) | 17 (24.6) |  |
| **Body mass index, kg/m<sup>2</sup>** | 27.6 ± 4.6 | 27.0 ± 4.3 | 27.5 ± 4.5 | 28.4 ± 4.8 | 27.8 ± 4.9 | 0.004 |
| **Depressive symptoms (0-30)** | 4.1 ± 3.5 | 3.2 ± 3.1 | 3.8 ± 3.3 | 5.2 ± 3.6 | 6.1 ± 4.3 | <0.001 |
| **Multimorbidity Index (0-11)** | 0.80 ± 0.86 | 0.72 ± 0.77 | 0.78 ± 0.85 | 0.93 ± 0.96 | 0.85 ± 0.96 | 0.081 |
| **Arthritis** | 492 (56.3) | 81 (31.8) | 197 (55.8) | 158 (79.8) | 56 (82.4) | <0.001 |
| **All prescription medications** | 4.4 ± 3.5 | 3.5 ± 2.9 | 4.2 ± 3.3 | 5.4 ± 3.6 | 6.0 ± 4.8 | <0.001 |
| Antidepressants | 168 (19.2) | 39 (15.2) | 57 (16.1) | 50 (25.3) | 22 (31.9) | <0.001 |
| Opioid analgesics | 16 (1.8) | 0 | 5 (1.4) | 6 (3.0) | 5 (7.2) | <0.001 |
| Nonsteroidal anti-inflammatories | 88 (10.0) | 7 (2.7) | 29 (8.2) | 36 (18.2) | 16 (23.2) | <0.001 |
| **400m walk time, sec** | 393.2 ± 72.1 | 373.5 ± 59.6 | 387.6 ± 61.9 | 415.3 ± 82.8 | 433.0 ± 96.1 | <0.001 |
| **Stair climbs time, sec** | 26.6 ± 7.2 | 25.9 ± 6.8 | 25.8 ± 6.6 | 28.1 ± 7.4 | 29.9 ± 9.4 | <0.001 |
| **Recalled pain measures** |  |  |  |  |  |  |
| **Global pain severity (0-10)** | 1.4 ± 1.5 | 0.0 ± 0.0 | 1.0 ± 0.5 | 2.7 ± 0.6 | 5.0 ± 0.8 | - |
| No pain site | 279 (31.7) | 226 (87.6) | 51 (14.4) | 1 (0.5) | 1 (1.4) | <0.001 |
| Single site pain | 291 (33.1) | 16 (6.2) | 167 (47.2) | 82 (41.4) | 26 (37.7) |  |
| Multisite pain | 240 (27.3) | 13 (5.0) | 117 (33.1) | 82 (41.4) | 28 (40.6) |  |
| Widespread pain | 69 (7.8) | 3 (1.2) | 19 (5.4) | 33 (16.7) | 14 (20.3) |  |
| **Somatosensory pain (0-22)** | 1.2 ± 2.8 | 0.5 ± 1.2 | 0.9 ± 2.1 | 1.8 ± 3.1 | 3.8 ± 5.8 | <0.001 |
| none (0) | 608 (70.2) | 203 (79.9) | 258 (73.9) | 115 (59.0) | 32 (47.1) | <0.001 |
| mild (>0-6) | 212 (24.5) | 49 (19.3) | 81 (23.2) | 60 (30.8) | 22 (32.4) |  |
| moderate-to-severe (>6) | 46 (5.3) | 2 (0.8) | 10 (2.9) | 20 (10.3) | 14 (20.6) |  |
| **Hip/knee stiffness (0-19)** | 2.5 ± 2.9 | 1.1 ± 1.8 | 2.3 ± 2.5 | 3.6 ± 3.3 | 5.4 ± 4.0 | <0.001 |
| **Lower body pain (0-51)** | 7.8 ± 6.8 | 3.4 ± 3.9 | 7.3 ± 5.5 | 11.4 ± 6.6 | 16.2 ± 8.4 | <0.001 |
| Lower back (0-12) | 2.3 ± 2.6 | 0.9 ± 1.7 | 2.1 ± 2.3 | 3.6 ± 2.6 | 5.1 ± 3.3 | <0.001 |
| Knees (0-13) | 2.4 ± 2.7 | 1.1 ± 1.8 | 2.5 ± 2.5 | 3.2 ± 2.8 | 4.4 ± 3.6 | <0.001 |
| Hips (0-13) | 1.4 ± 2.3 | 0.5 ± 1.2 | 1.3 ± 2.0 | 2.3 ± 2.8 | 3.2 ± 3.3 | <0.001 |
| Feet/ankles (0-13) | 1.6 ± 2.6 | 0.9 ± 1.8 | 1.4 ± 2.3 | 2.3 ± 2.9 | 3.5 ± 3.6 | <0.001 |
| **Movement-evoked pain measures** |  |  |  |  |  |  |
| **400m walk (-10 to +10)** | 1.0 ± 2.2 | 0.3 ± 1.3 | 0.6 ± 1.3 | 1.6 ± 2.7 | 3.8 ± 4.2 | <0.001 |
| Less pain upon movement (<0) | 29 (3.3) | 1 (0.4) | 6 (1.7) | 17 (8.6) | 5 (7.2) | <0.001 |
| No change (=0) | 578 (65.8) | 227 (88.3) | 251 (70.9) | 82 (41.4) | 18 (26.1) |  |
| More pain upon movement (>0) | 271 (30.9) | 29 (11.3) | 97 (27.4) | 99 (50.0) | 46 (66.7) |  |
| **Stair climb (-10 to +10)** | 0.1 ± 1.1 | 0.1 ± 0.9 | 0.0 ± 1.2 | 0.2 ± 1.3 | 0.2 ± 0.9 | 0.314 |
| Less pain upon movement (<0) | 36 (4.3) | 2 (0.8) | 16 (4.7) | 12 (6.3) | 6 (9.4) | 0.053 |
| No change (=0) | 725 (85.8) | 235 (94.8) | 296 (86.0) | 148 (78.3) | 46 (71.9) |  |
| More pain upon movement (>0) | 84 (9.9) | 11 (4.4) | 32 (9.3) | 29 (15.3) | 12 (18.8) |  |

**Notes for Table 1.**
Data shown as N (%), mean ± SD. *p* for linear trend across global pain severity (Brief Pain Inventory, short form; pain currently or in the past 24 hours) categories is calculated with linear regression models for those normally distributed variables and Jonckheere-Terpstra for skewed continuous or categorical data. Abbreviations: SOMMA, Study of Muscle, Mobility and Aging.

### Correlations of recalled and movement-evoked pain with physical performance

Global pain severity (BPI-*sf*) and lower body pain (lower back, hip, knee, foot/ankle) were moderately correlated with one another (*r*=0.57, *p*<0.001) (**Figure 1)**. Lower body pain was also correlated with hip/knee stiffness (*r*=0.57, *p*<0.001)—though participants did not always overlap in reporting both lower body pain and hip/knee stiffness (**Supplementary** Figure 1A). Somatosensory pain (NTSS-6) scores were weakly related to global pain severity (*r*=0.24, *p*<0.001) and lower body pain (*r*=0.31, *p*<0.001). Movement-evoked pain in the 400m walk or the stair climb tests were weakly correlated with one another (*r*=0.12, *p*<0.001) and to some extent, other recall pain measures. With exception to somatosensory pain (*r*=0.12, *p*<0.001), no other pain measures were significantly correlated with age.

**Figure 1.**
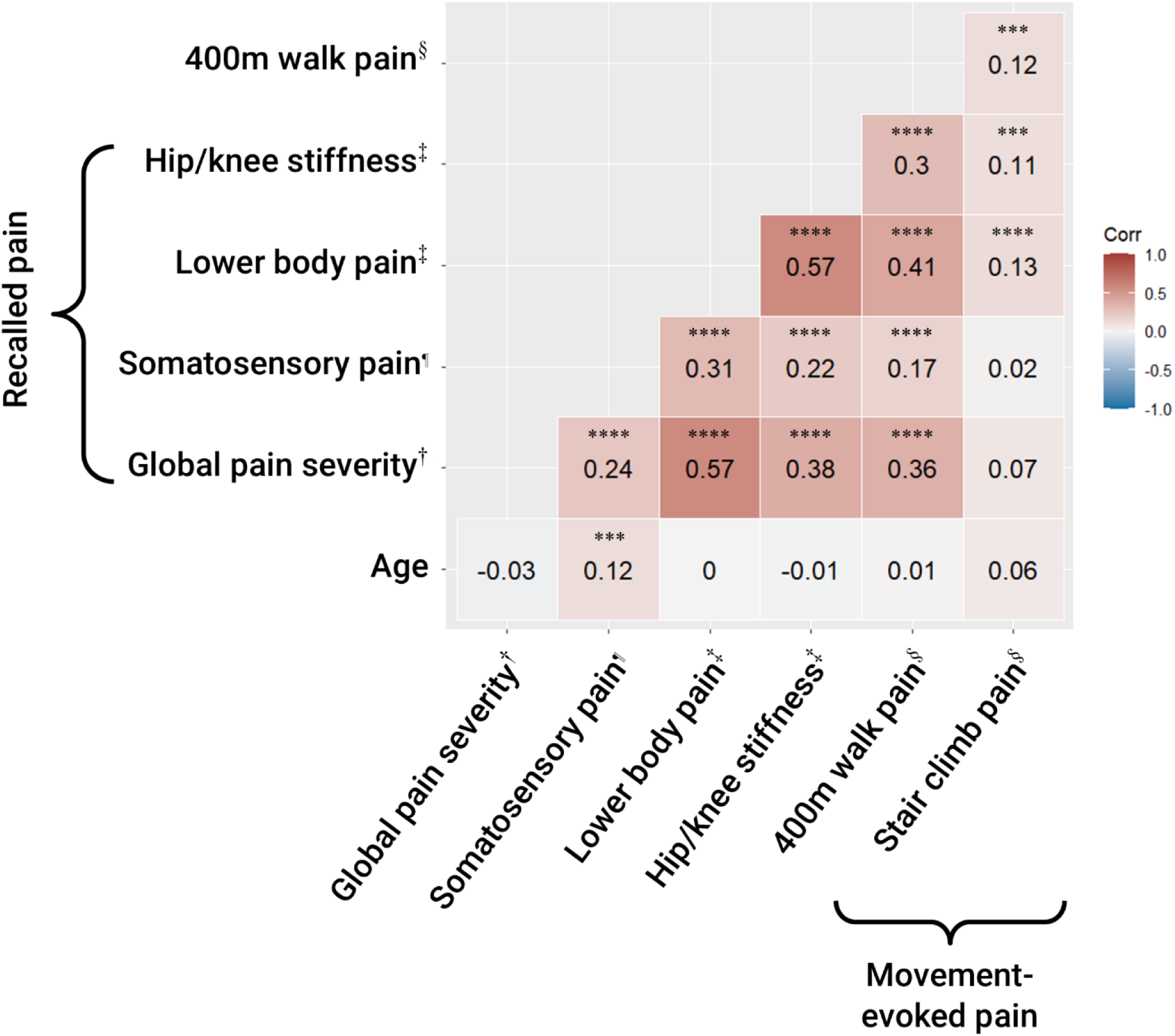
Correlations of recalled pain and movement-evoked pain measures. Figure 1 notations. † Global pain severity assessed by Brief Pain Inventory-short form (BPI-*sf*) on past 24-hour pain. ‡ Total lower body pain assessed by self-assessment questionnaire of pain severity and frequency (lower back, hips, knees, feet/ankles) and stiffness (hips or knees) within the past few months. § Movement-evoked pain was pain rating measured before, during, and after physical performance tests. ¶ Somatosensory pain was assessed by the Neuropathy Total Symptom Score, 6-item (NTSS-6) questionnaire on past 24-hour pain. Spearman’s correlation *p*-values are as follows: \**p*<0.05, \*\**p*<0.01, \*\*\**p*<0.001, \*\*\*\**p*<0.0001.

### The associations of recalled pain measures with physical performance

Greater levels of global pain severity, pain site classification, or somatosensory pain were all associated with longer 400m walk and repeat stair climb time (**Table 2** and **Supplementary Table 3**). On average, it takes 6.5 minutes (393 sec) for a participant to complete the 400m walk test and approximately 27 seconds to complete the repeat stair climb test. Single and multisite pain site classification were associated with longer 400m walk time when compared to those with no pain sites (*p*<0.010) (**Table 2**), and these associations were not significant for widespread pain (*p*=0.061) and all associations became attenuated in fully adjusted models except for single site pain (*p*<0.05). The associations of pain site with stair climb were not statistically significant. Moderate-to-severe (but not mild) somatosensory pain scores were associated with longer 400m walk time when compared to those with no somatosensory pain (*p*<0.001) (**Table 2**). The associations of somatosensory pain with the 400m walk time were independent of site, demographics (e.g., age, sex, race, ethnicity), and BMI, and these remained significant and were largely unattenuated when further adjusted for number of medications and depressive symptoms (*p*<0.001). For stair climb task, participants with moderate-to-severe (but not mild) somatosensory pain also had longer repeat stair climb times when compared to those with no somatosensory pain (*p*<0.05), and these associations became borderline significant in fully adjusted models (*p*=0.057). Of note, self-reported medical history of diabetes was not significant across greater somatosensory pain categories (**Supplementary Table 4**).

**Table 2.**
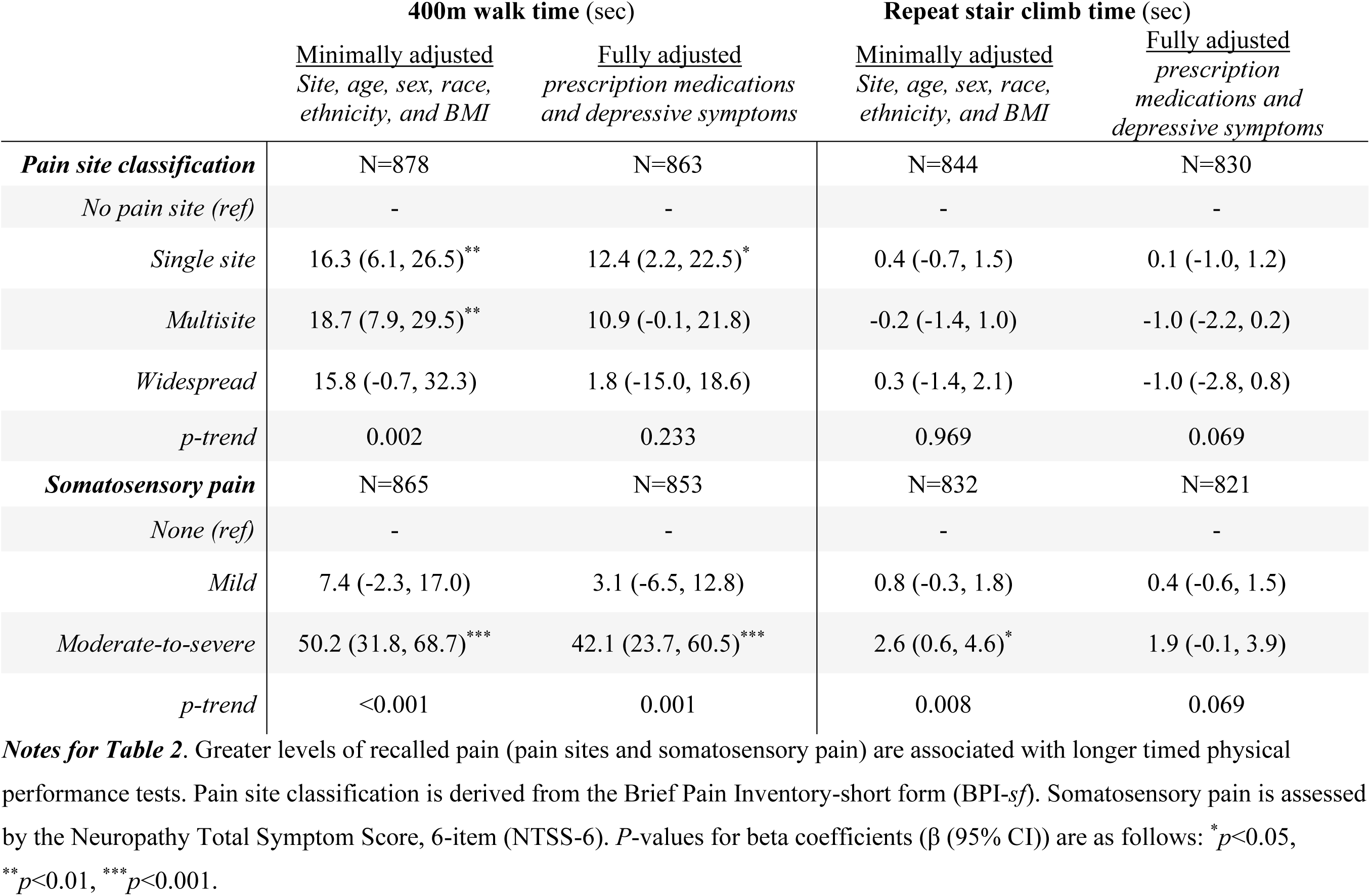
The associations of pain site classification and somatosensory pain with 400m walk and repeat stair climb tests.

|  | 400m walk time (sec) |  | Repeat stair climb time (sec) |  |
| --- | --- | --- | --- | --- |
|  | <u>Minimally adjusted</u><br>*Site, age, sex, race, ethnicity, and BMI* | <u>Fully adjusted</u><br>*prescription medications and depressive symptoms* | <u>Minimally adjusted</u><br>*Site, age, sex, race, ethnicity, and BMI* | <u>Fully adjusted</u><br>*prescription medications and depressive symptoms* |
| ***Pain site classification*** | N=878 | N=863 | N=844 | N=830 |
| *No pain site (ref)* | - | - | - | - |
| *Single site* | 16.3 (6.1, 26.5)** | 12.4 (2.2, 22.5)* | 0.4 (-0.7, 1.5) | 0.1 (-1.0, 1.2) |
| *Multisite* | 18.7 (7.9, 29.5)** | 10.9 (-0.1, 21.8) | -0.2 (-1.4, 1.0) | -1.0 (-2.2, 0.2) |
| *Widespread* | 15.8 (-0.7, 32.3) | 1.8 (-15.0, 18.6) | 0.3 (-1.4, 2.1) | -1.0 (-2.8, 0.8) |
| *p-trend* | 0.002 | 0.233 | 0.969 | 0.069 |
| ***Somatosensory pain*** | N=865 | N=853 | N=832 | N=821 |
| *None (ref)* | - | - | - | - |
| *Mild* | 7.4 (-2.3, 17.0) | 3.1 (-6.5, 12.8) | 0.8 (-0.3, 1.8) | 0.4 (-0.6, 1.5) |
| *Moderate-to-severe* | 50.2 (31.8, 68.7)*** | 42.1 (23.7, 60.5)*** | 2.6 (0.6, 4.6)* | 1.9 (-0.1, 3.9) |
| *p-trend* | <0.001 | 0.001 | 0.008 | 0.069 |

**Notes for Table 2.** Greater levels of recalled pain (pain sites and somatosensory pain) are associated with longer timed physical performance tests. Pain site classification is derived from the Brief Pain Inventory-short form (BPI-sf). Somatosensory pain is assessed by the Neuropathy Total Symptom Score, 6-item (NTSS-6). *P*-values for beta coefficients ( $\beta$ (95% CI)) are as follows: \* $p < 0.05$ , \*\* $p < 0.01$ , \*\*\* $p < 0.001$ .

Global pain severity was associated with longer 400m walk and stair climb time even when analyses were stratified by sex and knee osteoarthritis. Every one SD increment in global pain severity (BPI-*sf*) was associated with 16.8 seconds longer walk (*p*<0.001) and 1.1 seconds longer repeat stair climb (*p*<0.001) time (**Supplementary Table 3**); all these significant associations were retained in fully adjusted models (not shown). Sex differences in global pain severity were borderline significant with the 400m walk test (*p*-int=0.118). When stratified by women and men (not shown), every increment in global pain severity was associated with an additional 14.8 seconds (*p*<0.001) for women and 9.8 seconds (*p*<0.01) for men in 400m walk time, suggesting a stronger association in women than in men. Presence of any knee osteoarthritis may also modify the association of pain with performance on the 400m walk (*p*-int<0.04) and stair climb (*p*-int<0.10). In a subset of participants (N=658) with radiographic scoring on knee osteoarthritis (OA), sensitivity analyses (**Supplementary Table 3**) revealed that the association between pain and physical performance may be stronger in those with the worse knee OA status (400m walk, *p-int*=0.086; stair climb, *p-int*=0.148).

### Associations of recalled pain in specific anatomic sites with physical performance

Pain scores at specific anatomic sites—lower back, hip, knee, and foot/ankle were all significantly associated with slower 400m walk time (all, *p*<0.05) in separate models (**Table 3**). When all four anatomic pain scores were entered into one model, the association of hip pain with walk time was completely attenuated, while associations between the other three anatomic sites remained significant with walk time. For the 400m walk, every one SD increment of greater lower back pain was associated with the greatest time extended in both separate (β = 7.2 sec, *p*<0.01) and combined models (β = 4.9 sec, *p*<0.01). For the repeat stair climb, only knee pain was significantly associated with longer stair climb time (β = 0.9 sec, *p*<0.001). A higher composite score of lower body pain (summation of all four sites) was significantly associated with longer walk (β = 10.5 sec, *p*<0.001) and stair climb time (β = 0.6 sec, *p*<0.05). Greater hip and knee stiffness scores were similarly associated to longer walk (β = 9.4 sec, *p*<0.001) and stair climb (β = 1.0 sec, *p*<0.001) time. These significant associations were independent of age, sex, race-ethnicity, body mass index, total prescription medications, and depressive symptoms.

**Table 3.** Associations of recalled pain at specific anatomic sites with physical performance time in SOMMA older adults (≥70 years).

| Pain score<sup>†</sup> | N | 1 SD | Separate models<br>$\beta$ (95% CI) | One model<br>$\beta$ (95% CI) |
| --- | --- | --- | --- | --- |
| **400m walk time (sec.)** |  |  |  |  |
| *Lower back* | 863 | 2.6 | 7.2 (2.9, 11.5)** | 4.9 (0.2, 9.6)* |
| *Hips* | 862 | 2.3 | 5.4 (1.2, 9.7)* | 2.0 (-2.6, 6.6) |
| *Knees* | 863 | 2.7 | 6.4 (2.3, 10.6)** | 4.4 (0.1, 8.7)* |
| *Feet/Ankles* | 863 | 2.6 | 6.4 (2.2, 10.5)** | 4.6 (0.3, 8.8)* |
| *Total lower body pain* | 863 | 6.8 | 10.5 (6.1, 14.8)*** | - |
| *Hip/knee stiffness* | 863 | 2.9 | 9.4 (5.0, 13.7)*** | - |
| **Stair climb time (sec.)** |  |  |  |  |
| *Lower back* | 830 | 2.6 | 0.3 (-0.2, 0.7) | 0.2 (-0.3, 0.7) |
| *Hips* | 830 | 2.3 | 0.0 (-0.5, 0.4) | -0.3 (-0.8, 0.2) |
| *Knees* | 830 | 2.7 | 0.9 (0.5, 1.4)*** | 0.9 (0.5, 1.4)*** |
| *Feet/Ankles* | 830 | 2.6 | 0.2 (-0.2, 0.7) | 0.1 (-0.4, 0.5) |
| *Total lower body pain* | 830 | 6.8 | 0.6 (0.1, 1.1)* | - |
| *Hip/knee stiffness* | 830 | 2.9 | 1.0 (0.5, 1.5)*** | - |

**Notes for Table 3.**
<sup>†</sup>Pain scores were derived from self-assessment questionnaire items on severity and frequency of lower body pain and stiffness. 'Separate models': one model per row and one pain predictor per model. The 'One model' associations included all four pain scores (lower back, hips, knees, feet/ankles) into a singular model. *P*-values for linear regression outcomes are \**p*<0.05, \*\**p*<0.01, \*\*\**p*<0.001. All associations were adjusted for site, age, sex, race-ethnicity, body mass index, total number of prescription medications, and depressive symptoms (CESD-10; 10-item version of the Center of Epidemiologic Studies Depression Scale).

### Associations of movement-evoked pain with respective physical performance times

Movement-evoked pain scores were assessed separately for the 400m walk and repeat stair climb tests, and participants were grouped into either less pain upon movement, no change, and more pain upon movement. We found that greater movement-evoked pain during 400m walk and stair climb tests were separately associated with longer time to complete the respective physical performance task when compared to those with no change in pain (**Table 4**). Those with less pain (2.3 sec [-20.7,25.3]) had similar average 400m walk time as those with no change in pain (reference), but those with more pain (25.5 sec [16.5,34.6]) had at least an additional 25 seconds in average walk time when compared to those with no pain (*p-trend*<0.001). For the repeat stair climb test, there were similarly significant differences across those with no change in pain (reference), less pain (1.0 sec [-1.2,3.2]), and more pain (3.6 sec [2.1,5.1]) (*p-trend*<0.01). Fully adjusted associations of movement-evoked pain with corresponding physical performance time remained significant and largely unattenuated.

**Table 4.** Greater movement-evoked pain is associated with longer physical performance time in SOMMA older adults (≥70 years).

|  | <u>Minimally adjusted</u><br>*Site, age, sex, race,*<br>*ethnicity, and BMI* | <u>Fully adjusted</u><br>*prescription medications and*<br>*depressive symptoms* |
| --- | --- | --- |
| **400m walk time (sec.)** | N=877 | N=862 |
| Less pain upon movement | 2.3 (-20.7, 25.3) | 2.8 (-19.8, 25.5) |
| No change (*Ref*) | - | - |
| More pain upon movement | 25.5 (16.5, 34.6)*** | 19.5 (10.3, 28.7)*** |
| *p-trend* | <0.001 | <0.001 |
| **Repeat stair climb time (sec.)** | N=846 | N=832 |
| Less pain upon movement | 1.0 (-1.2, 3.2) | 0.6 (-1.5, 2.8) |
| No change (*Ref*) | - | - |
| More pain upon movement | 3.6 (2.1, 5.1)*** | 3.3 (1.9, 4.8)*** |
| *p-trend* | 0.001 | 0.001 |

***Notes for Table 4.***
Beta coefficients (95% CI) of physical performance time by groups of movement-evoked pain (less pain upon movement, no change, more pain upon movement). *P*-values for beta coefficients are as follows: \* $p < 0.05$ , \*\* $p < 0.01$ , \*\*\* $p < 0.001$ . Movement-evoked pain measures were assessed separately for the 400m walk (aggregate score) and repeat stair climb (delta score) tasks.

## Discussion

In this cross-sectional study of 879 older women and men, we used measures of recalled pain and movement-evoked pain to understand the associations of different pain measures with the 400m walk and repeat stair climb time. As expected, pain was common among SOMMA participants—nearly 70% reported at least one site of pain but pain parameters (site, severity, activity-related, sensation) were not homogenous across the participants. A greater degree of any pain measure–whether global or at an anatomic site, at rest or during movement, or sensory specific—were all robustly associated with worse physical performance. Given that different measures of recalled pain (global, lower body, hip/knee stiffness, somatosensory) and movement-evoked pain (during 400m walk and stair climb tests) are not strongly correlated with one another (*r*=0.2-0.6), these findings collectively suggest that various assessments within a study may be needed to detect pain in different individuals but may have similar associations with functional outcomes. Of note, the associations of pain site classification (i.e., no pain site, single site, multisite, widespread) with physical performance time were attenuated after fully adjusting for confounders. These findings are highly consistent with another cross-sectional study using a similar widespread definition.^2^ A longitudinal study^26^ demonstrated that both multisite and widespread pain predicted the risk of mobility difficulty and declines in physical performance. Overall, our findings show that both global and anatomic pain site specific measures are cross-sectionally associated with physical performance; the most robust pain-related measures associated with 400m walk and stair climb time in these older adults were global pain severity (BPI-*sf*), movement-evoked pain (aggregate score during the 400m walk), and somatosensory pain (NTSS-6). Pain measures less robustly associated with physical performance include pain site classification (i.e., none, single, multisite, widespread) and movement-evoked pain (delta score during the stair climb), and although total lower body pain and stiffness were weakly associated with stair climb time, these two pain measures were strongly associated with the 400m walk time.

### Recalled pain and movement-evoked pain in studies of aging and pain

One of the major strengths of this study is the inclusion of both recalled and movement-evoked pain assessments. When participants are asked to recall pain in questionnaires, they may or may not report pain that occurs during physical movement or pain that dissuades them to not move.^27^ This led us to consider that different pain measures may signal the presence of multiple influences or types of pain that are present in the aging population. For example, somatosensory pain and movement-evoked pain are weakly correlated with one another (*r*=0.17). Participants with the most severe movement-evoked pain (measured for 400m walk) or somatosensory pain (NTSS-6) took longer to complete the 400m walk. These results indicate that both pain assessments are relevant for identifying pain that is associated with worse physical performance. The inclusion of movement-evoked pain measures in aging studies is not yet standard,^28^ but our findings, among other studies,^29^ provide further support that these measures are important to include as they may capture pain that is distinct from measures of recalled pain.

### Recalled pain in specific anatomic sites and physical performance

Many studies of pain in aging have adapted questions from arthritis-related pain assessment tools.^30,31^ We refer to these measures as recalled pain in the lower body (lower back, hips, knees, feet/ankles) and hip/knee stiffness. When we consider pain measures at these four anatomic sites separately (4 models) and together (1 model) with physical performance, in general, composite pain and stiffness scores are more strongly related with physical performance than pain measures at singular anatomic sites. Among the anatomic sties, lower back pain and knee pain have the strongest associations with physical performance; these findings align with prior studies.^2,7,26^ We previously reported that pain and stiffness together are major determinants of walking speed.^14^ In this study we determined that greater hip/knee stiffness alone was significantly associated with physical performance, particularly repeat stair climb performance. These findings support the need for future studies to consider lower body pain and hip/knee stiffness separately to understand how each relates to physical function and its change over time.

### Recalled global pain was strongly associated with worse physical performance

Global pain severity (BPI-*sf*) is nonspecific to any anatomic site. This measure is associated with many other pain measures, in particular total lower body pain (*r*=0.57) and hip/knee stiffness (*r*=0.38). This may be because the most common pain sites reported by SOMMA participants included three of the four lower body sites, so the global pain severity may be a surrogate of lower body pain (and vice versa). Studies of pain in aging do not often use both global pain severity and lower body pain. In the few studies^10^ that have both measures, global pain severity (BPI-*sf*) was a stronger predictor of the risk of falls^7^ and mobility difficulty than anatomic site specific pain.^26^ Our cross-sectional results indicate that global pain severity may be one of the pain measures that is most strongly associated with both the 400m walk and repeat stair climb tests. In a sub cohort analysis (N=658), we found that the association between global pain severity and physical performance varied by knee OA status. This is unsurprising given that joint damage is the tenet of musculoskeletal pain, and 60% of the subset cohort had radiographic knee OA. In those with no knee OA, greater global pain severity remained significantly associated with longer 400m walk time. Greater global pain severity prevalence among those with knee OA reflects the known predisposition that those with knee OA tend to have greater and abnormal pain sensitization.^32^

### Recalled pain—somatosensory pain and physical performance

At baseline, almost one-third (29%) of SOMMA participants reported some somatosensory pain on the NTSS-6 questionnaire. Greater severity in somatosensory pain was strongly associated with worse physical performance. The NTSS-6 was originally developed in individuals afflicted with diabetic peripheral neuropathy^11,33,34^ to identify specific pain sensation symptoms. However, peripheral neuropathy also presents in non-diabetic older adults^35,36^ and may be more related to inflammatory nerve damage and sensitization shifts with advancing age.^37–40^ Notably, somatosensory pain is the only measure of pain that was significantly correlated with age (*r*=0.12) in this cohort of older adults over age 70, and self-reported history of diabetes was not significant across greater somatosensory pain categories. Future studies are warranted to determine the relationship of somatosensory pain (NTSS-6) scores with pain sensitivity assessments. Utilizing both somatosensory pain scores and measures of pain sensitivity may synergistically aid in the identification of peripheral nerve pain and other complex pain syndromes in older adults.

## Limitations

SOMMA enrollment criteria required participants to be able to complete the 400m walk within 15 minutes, which may have excluded those who have severe pain. For example, approximately 8% of SOMMA participants at baseline had widespread pain which is at least 5% lower than a different study of community-dwelling older adults that defined widespread similarly.^26^ Many older adults likely use over-the-counter pain medications instead of prescription medications to treat pain; we did not collect over the counter medications. Movement-evoked pain measures (e.g., 400m walk and stairs) will evoke pain in certain regions (e.g., knee, hip, sometimes low back but not always) and not others.^28^ Reporting of ‘no pain’ by global pain severity (BPI-*sf*) and ‘no pain’ by lower body pain was not completely congruent—this may be because the questionnaires were administered on different days. This discrepancy highlights the limitation of using recalled pain assessments in complex research studies. The SOMMA population is less racially and ethnically diverse than that of general US population, which presents two limitations. Our results may not generalize to under-represented groups, and we were unable to determine if associations varied across race and ethnic subgroups. Nonetheless, the strengths of the SOMMA cohort included extensive and detailed phenotyping of pain and gold standard measures of physical performance.

## Conclusions

Movement-evoked pain measures as well as measures of recalled pain specific to sensations or anatomic pain sites were all associated with physical performance. In addition, our results suggest that movement-evoked pain may increase identification of more severe forms of pain that lead to physical impairment. While it remains a challenge to parse out the different influences and origins of pain (e.g., neuropathic, nociceptive, nociplastic) based on pain measures, this work supports that future studies that use diverse pain assessments will improve our understanding of pain and its functional consequences during aging.

## Data Availability

All data produced are available online at https://sommaonline.ucsf.edu/, but users must agree to data use agreements.

## Acknowledgements

We thank all the SOMMA participants who enabled this research work, and we acknowledge all the staff and investigators listed.^15^ The manuscript was drafted by TM with formal analyses led by HNB. PMC, CBS, MH-R, SRB, ESS, and ABN provided the most critical edits and insight that led to substantial improvement of the manuscript. NEL, MCR, AAW, BHG, TLB, YC, and NWG reviewed multiple drafts of the manuscript. ABN, SRC, PMC, and SBK facilitated this work with funding acquisition, project administration, and study conceptualization. All authors reviewed and approved of the final manuscript.

---

**Supplementary Figure 1.**
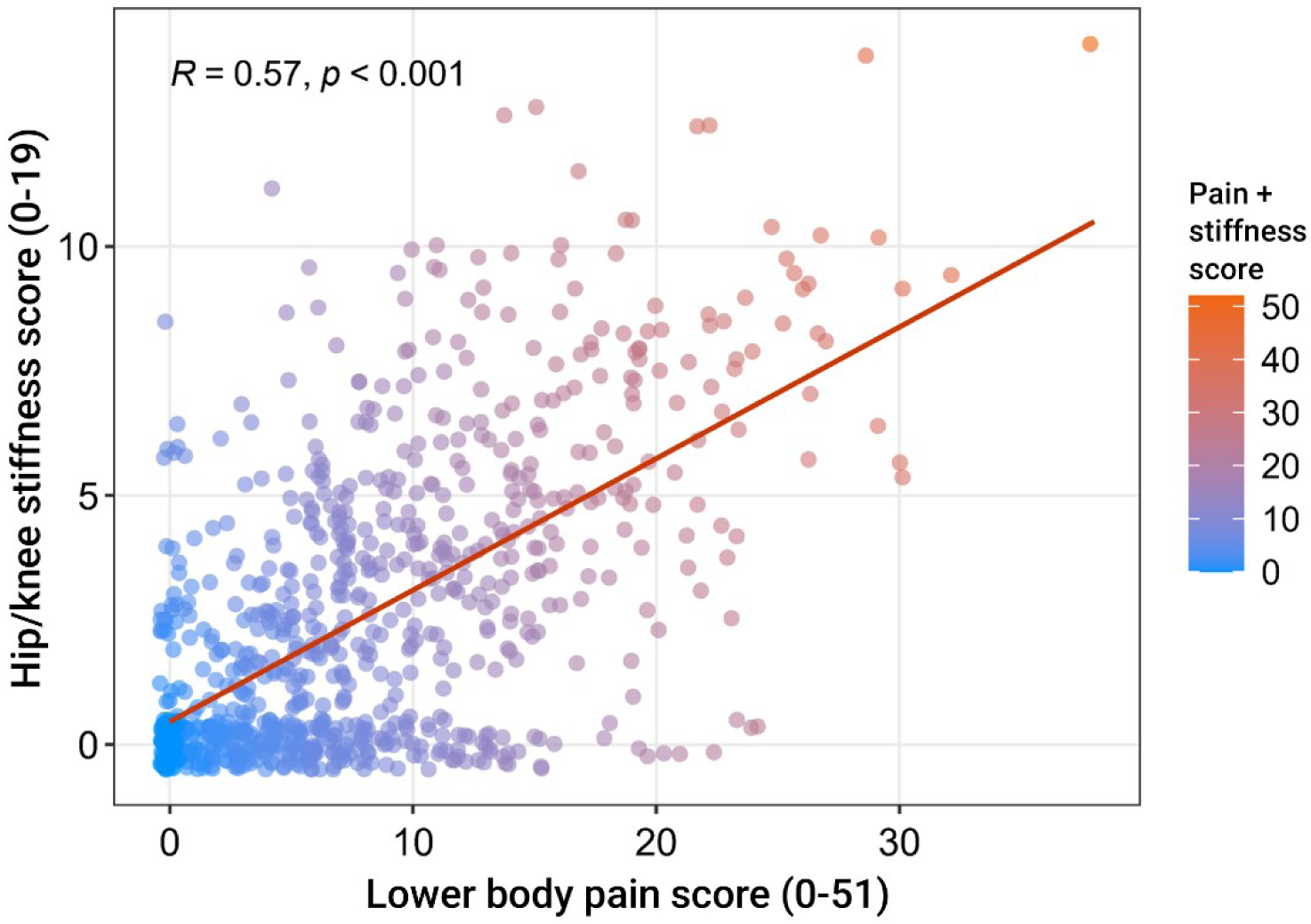
Spearman correlation of recalled measures of lower body pain and hip/knee stiffness.

**Supplementary Table 1.** Terminology for measures of pain in SOMMA.

| Pain measure |  |  | Assessment and definition |
| --- | --- | --- | --- |
| Recalled pain |  | *Global pain severity* | Brief pain inventory short form (BPI-*sf*) is a validated questionnaire that surveys global pain severity (**Table 1** stratified by this measure) in the past, current, worst, and average; **Supp. Table 3**. Anatomic site inventory (**Supp. Table 2**) and pain site classification (**Table 2**) uses a whole-body pain chart. |
|  | Anatomic site specific | *Somatosensory pain* | Neuropathy Total Symptom Score (NTSS-6) queries pain sensations (i.e., numbness, prickling, allodynia, lancinating, burning, aching) frequency and severity below the knees. **Table 2**. |
|  |  | *Lower body pain* | Pain questionnaire specific to lower back, hips, knees, and feet/ankles where pain intensity and frequency are rated for resting and/or physical activity related; **Table 3**. |
|  |  | *Hip/knee stiffness* | Stiffness in hips and knees were calculated separate from lower body pain; **Table 3**. |
| Movement-evoked pain |  | *Movement-evoked pain for the 400m walk test* | Pain was rated concurrently to the 400m walk test. Baseline pain was subtracted from summation of total pain during and after test (aggregate score); **Table 4**. |
|  |  | *Movement-evoked pain for the repeat stair climb test* | Pain was rated concurrently to the stair climb test. The difference between baseline and post-test pain was determined (delta score); **Table 4**. |

**Supplementary Table 2.** Pain inventory by anatomic site in SOMMA older adults (≥70 years).

|  | Overall<br>(N=879) | No pain<br>(N=279) | Single site<br>(N=318) | Multisite<br>(N=213) | Widespread<br>(N=69) |
| --- | --- | --- | --- | --- | --- |
| Lower back | 239 (27.19) | - | 90 (28.30) | 83 (38.97) | 66 (95.65) |
| Knees<sup>†</sup> | 188 (21.39) | - | 62 (19.50) | 88 (41.31) | 38 (55.07) |
| Shoulders<sup>†</sup> | 115 (13.08) | - | 33 (10.38) | 61 (28.64) | 21 (30.43) |
| Hips<sup>†</sup> | 105 (11.95) | - | 31 (9.75) | 50 (23.47) | 24 (34.78) |
| Other | 104 (11.83) | - | 29 (9.12) | 60 (28.17) | 15 (21.74) |
| Ankles/feet<sup>†</sup> | 85 (9.67) | - | 23 (7.23) | 44 (20.66) | 18 (26.09) |
| Legs<sup>†</sup> | 70 (7.96) | - | 16 (5.03) | 39 (18.31) | 15 (21.74) |
| Neck | 62 (7.05) | - | 11 (3.46) | 37 (17.37) | 14 (20.29) |
| Buttocks<sup>†</sup> | 33 (3.75) | - | 4 (1.26) | 19 (8.92) | 10 (14.49) |
| Mid-back | 32 (3.64) | - | 3 (0.94) | 17 (7.98) | 12 (17.39) |
| Upper back | 31 (3.53) | - | 5 (1.57) | 13 (6.10) | 13 (18.84) |
| Head | 21 (2.39) | - | 8 (2.52) | 10 (4.69) | 3 (4.35) |
| Abdomen | 13 (1.48) | - | 2 (0.63) | 11 (5.16) | 0 |
| Chest | 4 (0.46) | - | 1 (0.31) | 2 (0.94) | 1 (1.45) |

**Notes for Supplementary Table 2.**
Data shown as N (%). <sup>†</sup>Indicates pain laterality was simplified by counting any pain in left or right as a single count.

**Supplementary Table 3.** Global pain severity associations with physical performance and stratified by radiographic knee osteoarthritis in a subset of SOMMA participants.

| | N | 1 SD | $\beta$ (95% CI) |
| --- | --- | --- | --- |
| **400m walk time (sec.)** |  |  |  |
| All participants | 878 | 1.5 | 16.8 (12.7, 20.8)*** |
| Participant subset: | 658 |  |  |
| No knee OA | 262 | 1.4 | 10.4 (4.1, 16.7)** |
| Knee OA<sup>†</sup> | 153 | 1.5 | 16.7 (6.3, 27.0)** |
| Bilateral knee OA<sup>††</sup> | 243 | 1.5 | 19.2 (11.3, 27.1)*** |
| **Stair climb time (sec.)** |  |  |  |
| All participants | 844 | 1.5 | 1.1 (0.7, 1.6)*** |
| Participant subset: | 658 |  |  |
| No knee OA | 254 | 1.4 | 0.7 (0.0, 1.3) |
| Knee OA<sup>†</sup> | 139 | 1.5 | 1.3 (0.2, 2.5)* |
| Bilateral knee OA<sup>††</sup> | 234 | 1.5 | 1.4 (0.6, 2.2)** |

**Notes for Supplementary Table 3.**
In all participants (N=879) and a subset of participants (N=658) with radiographic scores on knee osteoarthritis (OA), all associations were adjusted for site, age, sex, race-ethnicity, and body mass index. *P*-values for linear regression outcomes are as follows: \**p*<0.05, \*\**p*<0.01, \*\*\**p*<0.001. <sup>†</sup>The presence of radiographic knee OA was defined as Kellgren-Lawrence (KLG) scores $\geq 2$ . <sup>††</sup>The presence of bilateral knee OA was defined as having KLG $\geq 2$ in both left and right sides in any combination of the tibio-femoral and patella-femoral sites.

**Supplementary Table 4.**
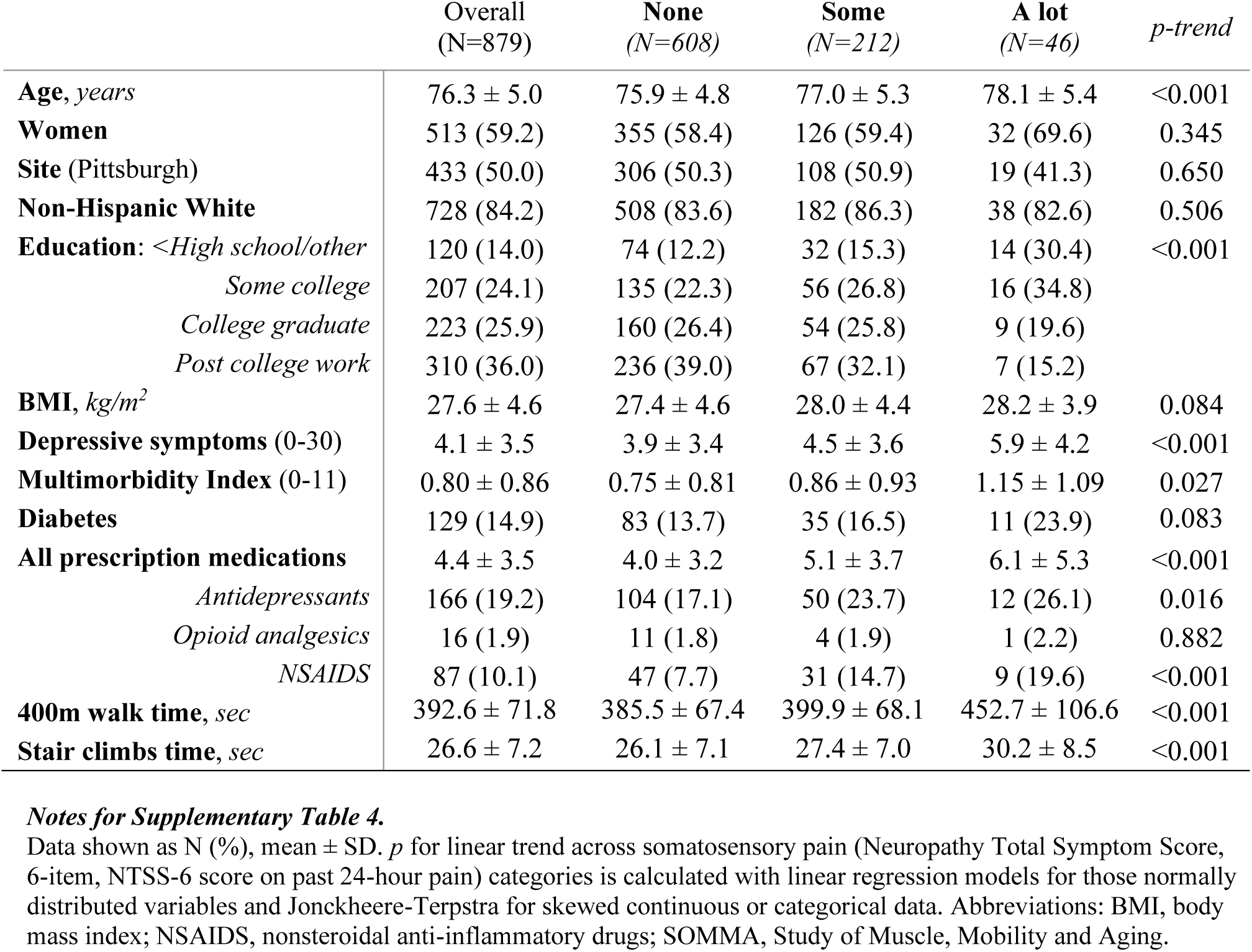
Baseline characteristics of SOMMA older adults (≥70 years) by somatosensory pain (NTSS-6).

|  | Overall<br>(N=879) | None<br>(N=608) | Some<br>(N=212) | A lot<br>(N=46) | *p-trend* |
| --- | --- | --- | --- | --- | --- |
| **Age, years** | 76.3 ± 5.0 | 75.9 ± 4.8 | 77.0 ± 5.3 | 78.1 ± 5.4 | <0.001 |
| **Women** | 513 (59.2) | 355 (58.4) | 126 (59.4) | 32 (69.6) | 0.345 |
| **Site (Pittsburgh)** | 433 (50.0) | 306 (50.3) | 108 (50.9) | 19 (41.3) | 0.650 |
| **Non-Hispanic White** | 728 (84.2) | 508 (83.6) | 182 (86.3) | 38 (82.6) | 0.506 |
| **Education:** <High school/other | 120 (14.0) | 74 (12.2) | 32 (15.3) | 14 (30.4) | <0.001 |
| Some college | 207 (24.1) | 135 (22.3) | 56 (26.8) | 16 (34.8) |  |
| College graduate | 223 (25.9) | 160 (26.4) | 54 (25.8) | 9 (19.6) |  |
| Post college work | 310 (36.0) | 236 (39.0) | 67 (32.1) | 7 (15.2) |  |
| **BMI, kg/m<sup>2</sup>** | 27.6 ± 4.6 | 27.4 ± 4.6 | 28.0 ± 4.4 | 28.2 ± 3.9 | 0.084 |
| **Depressive symptoms (0-30)** | 4.1 ± 3.5 | 3.9 ± 3.4 | 4.5 ± 3.6 | 5.9 ± 4.2 | <0.001 |
| **Multimorbidity Index (0-11)** | 0.80 ± 0.86 | 0.75 ± 0.81 | 0.86 ± 0.93 | 1.15 ± 1.09 | 0.027 |
| **Diabetes** | 129 (14.9) | 83 (13.7) | 35 (16.5) | 11 (23.9) | 0.083 |
| **All prescription medications** | 4.4 ± 3.5 | 4.0 ± 3.2 | 5.1 ± 3.7 | 6.1 ± 5.3 | <0.001 |
| Antidepressants | 166 (19.2) | 104 (17.1) | 50 (23.7) | 12 (26.1) | 0.016 |
| Opioid analgesics | 16 (1.9) | 11 (1.8) | 4 (1.9) | 1 (2.2) | 0.882 |
| NSAIDS | 87 (10.1) | 47 (7.7) | 31 (14.7) | 9 (19.6) | <0.001 |
| **400m walk time, sec** | 392.6 ± 71.8 | 385.5 ± 67.4 | 399.9 ± 68.1 | 452.7 ± 106.6 | <0.001 |
| **Stair climbs time, sec** | 26.6 ± 7.2 | 26.1 ± 7.1 | 27.4 ± 7.0 | 30.2 ± 8.5 | <0.001 |

**Notes for Supplementary Table 4.**
Data shown as N (%), mean ± SD. *p* for linear trend across somatosensory pain (Neuropathy Total Symptom Score, 6-item, NTSS-6 score on past 24-hour pain) categories is calculated with linear regression models for those normally distributed variables and Jonckheere-Terpstra for skewed continuous or categorical data. Abbreviations: BMI, body mass index; NSAIDS, nonsteroidal anti-inflammatory drugs; SOMMA, Study of Muscle, Mobility and Aging.

## References

1. Patel K V., Guralnik JM, Dansie EJ, Turk DC. Prevalence and impact of pain among older adults in the United States: Findings from the 2011 National Health and Aging Trends Study. Pain. 2013;154(12):2649–2657. doi:10.1016/j.pain.2013.07.029

2. Eggermont LHP, Bean JF, Guralnik JM, Leveille SG. Comparing pain severity versus pain location in the MOBILIZE Boston study: Chronic pain and lower extremity function. Journals Gerontol - Ser A Biol Sci Med Sci. 2009;64(7):763–770. doi:10.1093/gerona/glp016

3. Ettinger WH, Fried LP, Harris T, et al. Self-Reported Causes of Physical Disability in Older People: The Cardiovascular Health Study. J Am Geriatr Soc. 1994;42(10):1035–1044. doi:10.1111/j.1532-5415.1994.tb06206.x

4. Patel K V., Phelan EA, Leveille SG, et al. High prevalence of falls, fear of falling, and impaired balance in older adults with pain in the United States: Findings from the 2011 National Health and Aging Trends Study. J Am Geriatr Soc. 2014;62(10):1844–1852. doi:10.1111/jgs.13072

5. Cai Y, Leveille SG, Shi L, Chen P, You T. Chronic pain and risk of injurious falls in community-dwelling older adults. Journals Gerontol - Ser A Biol Sci Med Sci. 2021;76(9):E179–E186. doi:10.1093/gerona/glaa249

6. Eggermont LHP, Penninx BWJH, Jones RN, Leveille SG. Depressive symptoms, chronic pain, and falls in older community-dwelling adults: The mobilize Boston study. J Am Geriatr Soc. 2012;60(2):230–237. doi:10.1111/j.1532-5415.2011.03829.x

7. Leveille SG, Jones RN, Kiely DK, et al. Chronic musculoskeletal pain and the occurrence of falls in an older population. Jama. 2009;302(20):2214–2221. doi:10.1001/jama.2009.1738

8. Fitzcharles MA, Cohen SP, Clauw DJ, Littlejohn G, Usui C, Häuser W. Nociplastic pain: towards an understanding of prevalent pain conditions. Lancet. 2021;397(10289):2098–2110. doi:10.1016/S0140-6736(21)00392-5

9. Dubin AE, Patapoutian A. Nociceptors: the sensors of the pain pathway. J Clin Invest. 2010;120(11):3760–3772. doi:10.1172/JCI42843

10. Mendoza T, Mayne T, Rublee D, Cleeland C. Reliability and validity of a modified Brief Pain Inventory short form in patients with osteoarthritis. Eur J Pain. 2006;10(4):353. doi:10.1016/j.ejpain.2005.06.002

11. Bastyr EJ, Price KL, Bril V. Development and validity testing of the neuropathy total symptom score-6: Questionnaire for the study of sensory symptoms of diabetic peripheral neuropathy. Clin Ther. 2005;27(8):1278–1294. doi:10.1016/j.clinthera.2005.08.002

12. Wolfe F. Determinants of WOMAC function, pain and stiffness scores: Evidence for the role of low back pain, symptom counts, fatigue and depression in osteoarthritis, rheumatoid arthritis and fibromyalgia. Rheumatology. 1999;38(4):355–361. doi:10.1093/rheumatology/38.4.355

13. Wideman TH, Finan PH, Edwards RR, et al. Increased sensitivity to physical activity among individuals with knee osteoarthritis: Relation to pain outcomes, psychological factors, and responses to quantitative sensory testing. Pain. 2014;155(4):703–711. doi:10.1016/j.pain.2013.12.028

14. Cummings SR, Lui LY, Glynn NW, et al. Energetics and clinical factors for the time required to walk 400 m: The Study of Muscle, Mobility and Aging (SOMMA). J Am Geriatr Soc. Published online 2024. doi:10.1111/jgs.18763

15. Cummings SR, Newman AB, Coen PM, et al. The Study of Muscle, Mobility and Aging (SOMMA): A Unique Cohort Study About the Cellular Biology of Aging and Age-related Loss of Mobility. Journals Gerontol Ser A Biol Sci Med Sci. 2023;78(11):2083–2093. doi:10.1093/gerona/glad052

16. Irwin M, Artin KH, Oxman MN. Screening for depression in the older adult: Criterion validity of the 10-item Center for Epidemiological Studies Depression Scale (CES-D). Arch Intern Med. 1999;159(15):1701–1704. doi:10.1001/archinte.159.15.1701

17. Distefano G, Harrison S, Lynch J, et al. Skeletal muscle composition, power, and mitochondrial energetics in older men and women with knee osteoarthritis. Arthritis Rheumatol. 2024;0(0):1–11. doi:10.1002/art.42953

18. Kellgren JH, Lawrence JS. Radiological assessment of osteo-arthrosis. Ann Rheum Dis. 1957;16(4):494–502. doi:10.1136/ard.16.4.494

19. Fung KW, McDonald C, Bray BE. RxTerms - a drug interface terminology derived from RxNorm. AMIA Annu Symp Proc. 2008;2008:227–231. Accessed May 16, 2024. /pmc/articles/PMC2655997/

20. Bodenreider O, Peters L. RxMix – Use of NLM drug APIs by non-programmers. Published online 2017:2291. Accessed May 16, 2024. www.amia.org

21. Lagerlund O, Strese S, Fladvad M, Lindquist M. WHODrug: A Global, Validated and Updated Dictionary for Medicinal Information. Ther Innov Regul Sci. 2020;54(5):1116–1122. doi:10.1007/s43441-020-00130-6

22. Cleeland CS. The Brief Pain Inventory User Guide. Published online 2009:1–66. Accessed May 13, 2024. www.mdanderson.org

23. Wolfe F, Smythe HA, Yunus MB, et al. The american college of rheumatology 1990 criteria for the classification of fibromyalgia. Arthritis Rheum. 1990;33(2):160–172. doi:10.1002/art.1780330203

24. Simonsick EM, Montgomery PS, Newman AB, Bauer DC, Harris T. Measuring fitness in healthy older adults: The health ABC long distance corridor walk. J Am Geriatr Soc. 2001;49(11):1544–1548. doi:10.1046/j.1532-5415.2001.4911247.x

25. Lange-Maia BS, Newman AB, Strotmeyer ES, Harris TB, Caserotti P, Glynn NW. Performance on fast- and usual-paced 400-m walk tests in older adults: are they comparable? Aging Clin Exp Res. 2015;27(3):309–314. doi:10.1007/s40520-014-0287-y

26. Eggermont LHP, Leveille SG, Shi L, et al. Pain characteristics associated with the onset of disability in older adults: The maintenance of balance, independent living, intellect, and zest in the elderly boston study. J Am Geriatr Soc. 2014;62(6):1007–1016. doi:10.1111/jgs.12848

27. Knox PJ, Simon CB, Pohlig RT, et al. Construct validity of movement-evoked pain operational definitions in older adults with chronic low back pain. Pain Med (United States). 2023;24(8):985–992. doi:10.1093/pm/pnad034

28. Simon CB, Hicks GE, Pieper CF, Byers Kraus V, Keefe FJ, Colón-Emeric C. A Novel Movement-Evoked Pain Provocation Test for Older Adults With Persistent Low Back Pain: Safety, Feasibility, and Associations With Self-reported Physical Function and Usual Gait Speed. Clin J Pain. 2023;39(4):166–174. doi:10.1097/AJP.0000000000001101

29. Butera KA, Chimenti RL, Alsouhibani AM, et al. Through the Lens of Movement-Evoked Pain: A Theoretical Framework of the “Pain-Movement Interface” to Guide Research and Clinical Care for Musculoskeletal Pain Conditions. J Pain. 2024;25(7):104486. doi:10.1016/j.jpain.2024.01.351

30. Thakral M, Shi L, Foust JB, et al. Persistent pain quality as a novel approach to assessing risk for disability in community-dwelling elders with chronic pain. Journals Gerontol - Ser A Biol Sci Med Sci. 2019;74(5):733–741. doi:10.1093/gerona/gly133

31. Ogawa EF, Shi L, Bean JF, et al. Chronic Pain Characteristics and Gait in Older Adults: The MOBILIZE Boston Study II. Arch Phys Med Rehabil. 2020;101(3):418–425. doi:10.1016/j.apmr.2019.09.010

32. Rice D, Nijs J, Kosek E, et al. Exercise-Induced Hypoalgesia in Pain-Free and Chronic Pain Populations: State of the Art and Future Directions. J Pain. 2019;20(11):1249–1266. doi:10.1016/j.jpain.2019.03.005

33. Vinik AI, Bril V, Kempler P, et al. Treatment of symptomatic diabetic peripheral neuropathy with the protein kinase C β-inhibitor ruboxistaurin mesylate during a 1-year, randomized, placebo-controlled, double-blind clinical trial. Clin Ther. 2005;27(8):1164–1180. doi:10.1016/j.clinthera.2005.08.001

34. Tesfaye S, Tandan R, Bastyr EJ, Kles KA, Skljarevski V, Price KL. Factors that impact symptomatic diabetic peripheral neuropathy in placebo-administered patients from two 1-year clinical trials. Diabetes Care. 2007;30(10):2626–2632. doi:10.2337/dc07-0608

35. Baldereschi M, Inzitari M, Carlo A Di, Farchi G, Scafato E, Inzitari D. Epidemiology of distal symmetrical neuropathies in the Italian elderly. Neurology. Published online 2007. Accessed July 24, 2024. https://www.neurology.org

36. Hicks CW, Wang D, Windham BG, Matsushita K, Selvin E. Prevalence of peripheral neuropathy defined by monofilament insensitivity in middle-aged and older adults in two US cohorts. Sci Rep. 2021;11(1):1–11. doi:10.1038/s41598-021-98565-w

37. Ward RE, Caserotti P, Cauley JA, et al. Mobility-related consequences of reduced lower-extremity peripheral nerve function with age: A systematic review. Aging Dis. 2016;7(4):466–478. doi:10.14336/AD.2015.1127

38. Lange-Maia BS, Cauley JA, Newman AB, et al. Sensorimotor peripheral nerve function and physical activity in older men. J Aging Phys Act. 2016;24(4):559–566. doi:10.1123/japa.2015-0207

39. Lange-Maia BS, Newman AB, Jakicic JM, et al. Relationship between sensorimotor peripheral nerve function and indicators of cardiovascular autonomic function in older adults from the Health, Aging and Body Composition Study. Exp Gerontol. 2017;96:38–45. doi:10.1016/j.exger.2017.04.007

40. Doshi S, Moorthi RN, Fried LF, et al. Chronic kidney disease as a risk factor for peripheral nerve impairment in older adults: A longitudinal analysis of Health, Aging and Body Composition (Health ABC) study. PLoS One. 2020;15(12 December). doi:10.1371/journal.pone.0242406

